# Regularity in occurrence of respiratory-related events in sleep predicts cardiovascular disease and mortality

**DOI:** 10.64898/2026.02.25.26347037

**Authors:** Angela J. Senders, Ali Azarbarzin, Farhad Kaffashi, Kenneth A. Loparo, Susan Redline, Matthew P. Butler

## Abstract

**Background:** Obstructive sleep apnea (OSA), as measured by the Apnea Hypopnea Index (AHI), is associated with adverse outcomes. Measures that characterize the temporal variability in events may provide information over and beyond a simple summary of event frequency as measured by the AHI.

**Research Question:** To assess whether temporal variability in the occurrence of obstructive apnea/hypopneas during the night is associated with all-cause mortality or incident cardiovascular disease (CVD).

**Study Design and Methods:** Data from the Sleep Heart Health Study (SHHS), a prospective multi-site community-based cohort were analyzed. For each person, the intervals between apnea/hypopnea events (inter-event interval; IEI) were used to calculate a coefficient of variation for their IEIs (IEI_CV). Risk for mortality (n=5,701) and incident CVD (n=4,373) were estimated by adjusted Cox proportional hazard models. Sensitivity analyses were conducted to test potential explanatory variables such as hypoxic burden and duration of uninterrupted sleep.

**Results:** In 11.8 years of follow-up (median, IQR 10.6-12.2), 1,287 deaths occurred. After adjusting for potential confounders, including OSA severity, participants in the lowest quartile of IEI_CV (Q1) had a 40% higher risk of all-cause mortality compared with those in the highest quartile (Q4) (hazard ratio [HR] = 1.40; 95% confidence interval [CI], 1.20-1.64). In 11.5 years of follow-up (IQR 7.9-12.7), 867 CVD events occurred. The adjusted hazard rate for CVD was 29% higher (HR=1.29 [1.06-1.56]) for those with less variable IEI. Minimal reductions in effects sizes were observed after additional adjustment for hypoxic burden and additional novel and traditional covariates. In sensitivity analyses, adjusting for the longest bout of uninterrupted sleep without respiratory events attenuated the association for CVD incidence (HR=1.15 [0.89-1.50]).

**Interpretation:** The temporal distribution of respiratory events - specifically, less variability in inter-event intervals (more regular event occurrences) - is associated with higher mortality and incident CVD.

## INTRODUCTION

Obstructive sleep apnea (OSA) is associated with increased cardiovascular disease (CVD) risk and mortality, though the features of OSA that best carry prognostic value are still not well understood.^1,2^ Clinically, OSA is diagnosed by the frequency of obstructive events (apnea hypopnea index, AHI), but this metric has been repeatedly criticized for its low prognostic value for long term outcomes and its inability to distinguish among OSA subtypes that reflect differences in mechanistic traits (endophenotypes) and downstream consequences.^3–6^ One shortcoming of the AHI is that it defines severity based only on the number of events, and not on characteristics that may better indicate pathophysiological pathways.^7–11^

Physiological consequences of OSA may arise not only from the frequency of events, but also from their timing and clustering. Normal respiration during sleep varies in rate and depth, reflecting changes in sleep stage, metabolic demand, and the respiratory variability related to maintaining normal lung function.^12–16^ OSA disrupts these normal nocturnal breathing patterns, but the nature of this disruption varies substantially across the night and across people. Obstructive events are not uniformly distributed, with their occurrence and duration influenced by body position, sleep stage, and time of night.^8,17–22^ As a result, temporal patterns of obstructive events can range from highly clustered to more regularly spaced, yet these patterns have received relatively little attention. In other physiological systems, variability—such as in heart rate or arousal dynamics^23,24^—is often considered a marker of better health, but whether variability in obstructive respiratory events across the sleep period has prognostic value is not known. Greater variability in event timing necessarily produces intervals of both high and low event frequency: periods of clustering may amplify arousal-related autonomic and hypoxemic stress, whereas longer inter-event intervals may allow greater restorative sleep.

To test the hypothesis that variability in respiratory event timing across the sleep period provides independent prognostic information for CVD and mortality, we conducted a secondary analysis of data from the Sleep Heart Health Study. We quantified variability in inter-event intervals (IEI) and examined its associations with incident CVD and all-cause mortality. In addition, we evaluated a measure of sleep continuity to explore its potential to explain associations between IEI variability and adverse outcomes.

## METHODS

### Study Design

The Sleep Heart Health Study (SHHS) is a prospective multi-center cohort (N=6,441) designed to investigate associations between sleep-disordered breathing and health outcomes.^25^ The baseline visit for SHHS consisted of sleep questionnaires and an overnight polysomnography. Parent cohorts provided covariate data and surveilled incident CVD events until 2011. Data here are from five of the original cohorts (N=5,804), publicly available from the National Sleep Research Resource (NSRR; https://sleepdata.org).^26^ The Institutional Review Board of Oregon Health & Science University determined that no IRB oversight was needed because the data are pseudonymized.

### Sample Characteristics

For the mortality endpoint, data were drawn from the Cardiovascular Health Study, Atherosclerosis Risk in Communities Study, Framingham Heart Study, and the New York and Tucson cohorts of SHHS. The study sample included 5,701 participants who were observed for 62,785 person-years; median follow-up time was 11.8 years (IQR 10.6-12.8 years) (**Figure 1A**).

**Figure 1.**
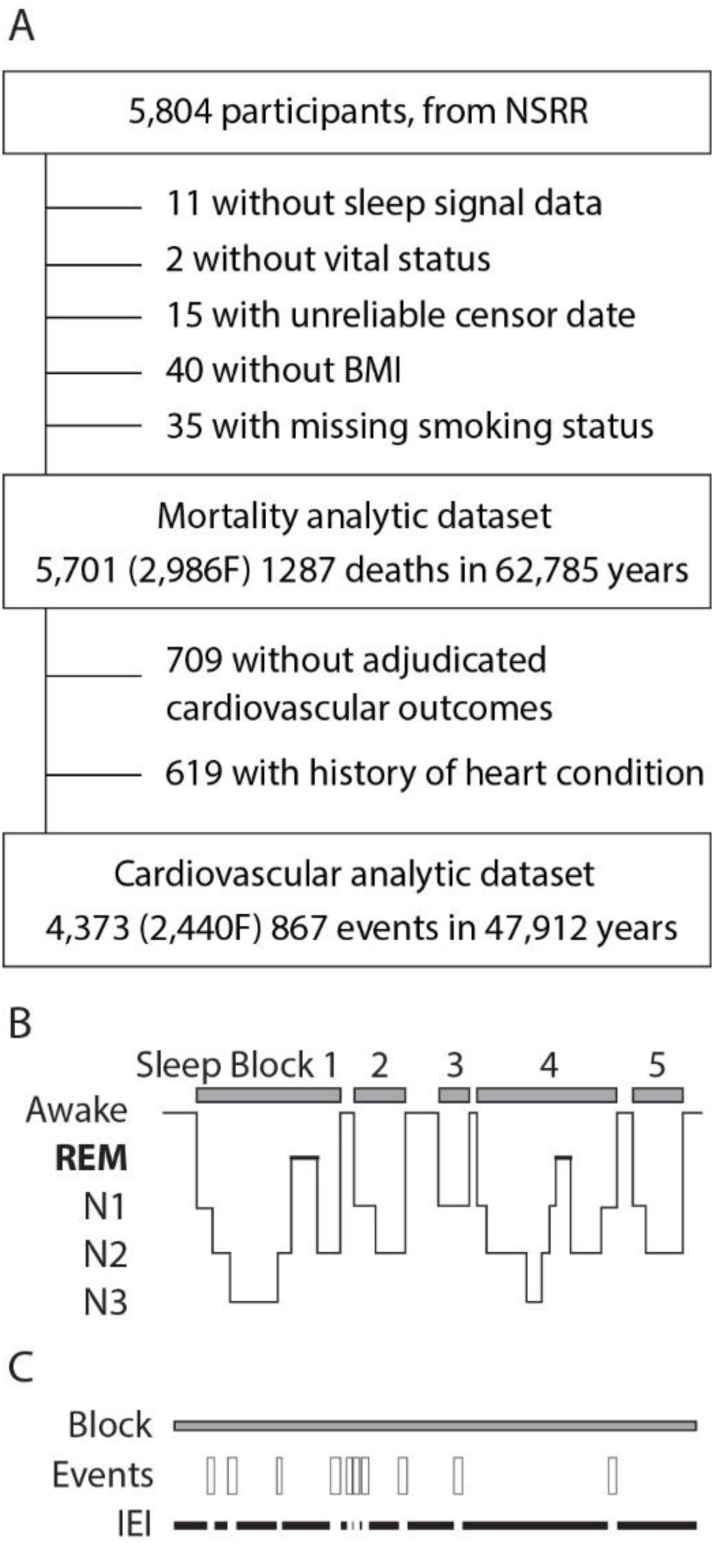
A) Derivation of the analytic study samples for mortality and incident CVD endpoints. The 709 participants without adjudicated cardiovascular outcomes are those in the New York parent cohort. B) Schematic showing the division of sleep into blocks and C) the IEIs within a block.

For the CVD endpoint, we further excluded those with a history of congestive heart failure, coronary heart disease, or stroke at baseline (n=619), as well as all participants with missing CVD data (n=709; the New York cohort was excluded due to inadequate follow-up CVD data). The final sample included 4,373 participants who were observed for 47,911 person-years; median follow-up time was 11.4 years (IQR 8.6-12.4 years).

### Independent Variable: Inter-event Interval Variability

Scored polysomnograms in XML were acquired from the NSRR. Annotated obstructive respiratory events (apneas and hypopneas) were defined by reductions in amplitude of the thermistry or inductance plethysmography signals for at least 10 sec. Two measures of event frequency were used: 1) AHI3 includes obstructive events that are each accompanied by ≥3% desaturation, and 2) AHI0 includes obstructive events regardless of desaturation criteria.

For each participant, inter-event intervals (IEI) were measured using all manually annotated obstructive apneas and hypopneas (no central events). First, the overnight polysomnogram was divided into blocks of continuous sleep; blocks were separated by at least one 30-sec epoch of wakefulness. Then, within each block of sleep, IEIs were measured: 1) from the start of the block to the first respiratory event, 2) from the end of a respiratory event to the onset of the next event, and 3) from the end of an event to the end of the sleep block (**Figure 1B**). For each individual, IEI variability was calculated as the IEI coefficient of variation (IEI_CV, standard deviation divided by their mean IEI). This corrects for mean IEI, and by extension, for the frequency of apneas and hypopnea (i.e., AHI0). IEI_CV was categorized into quartiles; quartile 4 (highest IEI variability) served as the reference category.

### Dependent Variables: Mortality and CVD

All-cause mortality was surveilled between enrollment (1995-1998) to 2011 as previously described^27^ Incident CVD was defined as the first event of fatal or non-fatal myocardial infarction, stroke, congestive heart failure, or revascularization procedure (angioplasty, coronary stent placement, or coronary artery bypass grafting). Ascertainment and confirmation of CVD events were conducted by the parent studies according to cohort-specific protocols.^25,28^

### Covariates

The base model included baseline values of: age (continuous), sex (male/female), BMI (under/normal, overweight, or obese), smoking status (non/current/former), race (self-reported as White/Black/other), OSA clinical severity (mild/moderate/severe OSA defined by AHI3 thresholds of 5, 15, or 30 events/hour), hypertension (yes/no, based on baseline BP or medication), diabetes (yes/no, based on self-report or use of medications), and parent cohort. In sensitivity analyses, additional covariates were prevalent heart disease (yes/no, for the all-cause mortality analyses only) and several newer metrics of OSA severity: (1) average duration of events (quartiles)^8^, (2) the mean heart rate response to events (ΔHR, maximum HR post-event maximum HR – intra-event minimum HR), modeled categorically using established cut points, given that both low and high ΔHR are associated with increased risk (ΔHR categories: low <5.8; mid 5.8–10.1; high >10.1),^29^ and (3) hypoxic burden, defined as the cumulative area under desaturation curve per hour of sleep, as described previously.^9^ See **e-Table 1** for variables used, sources, and definitions of prevalent conditions.

**Table 1.**
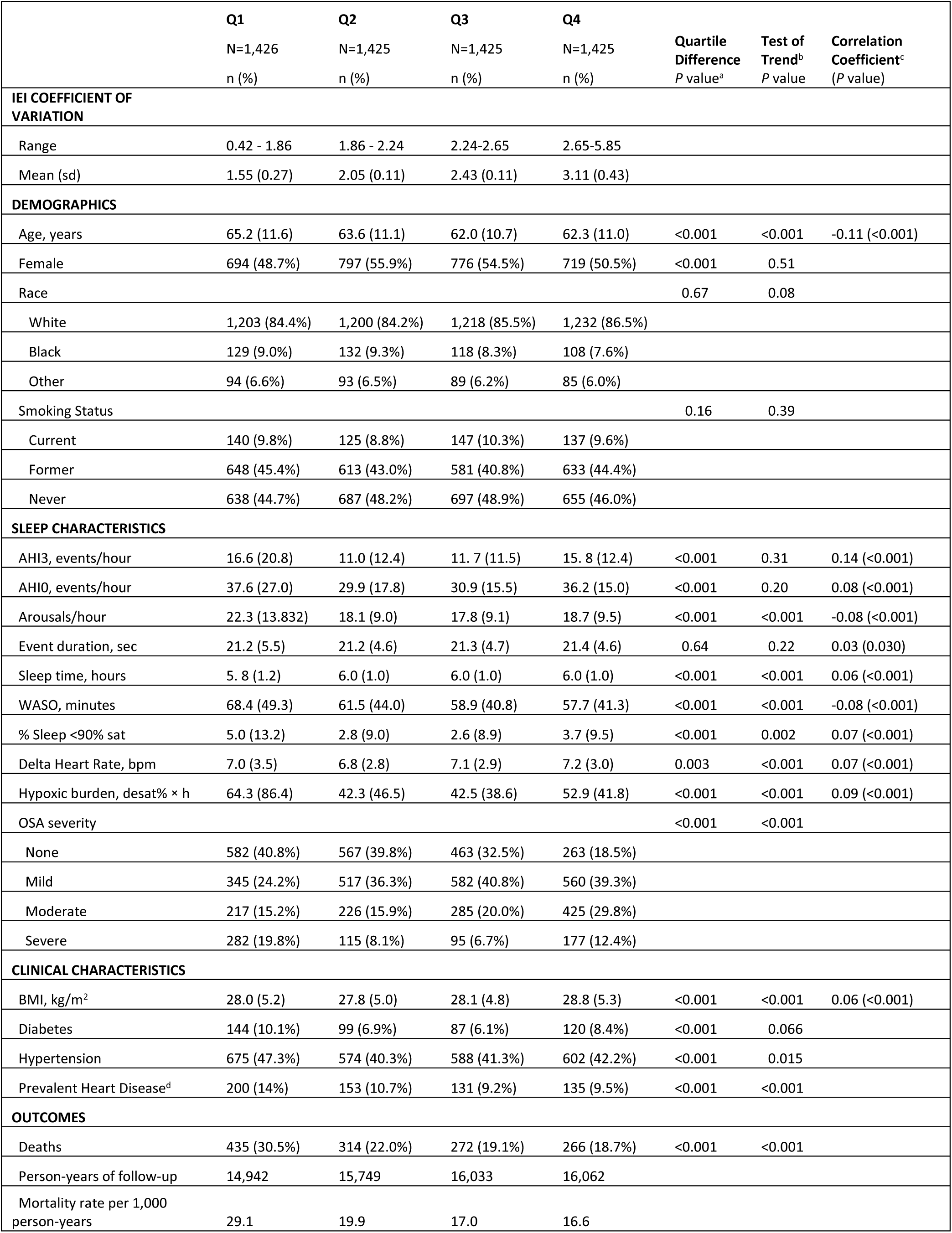
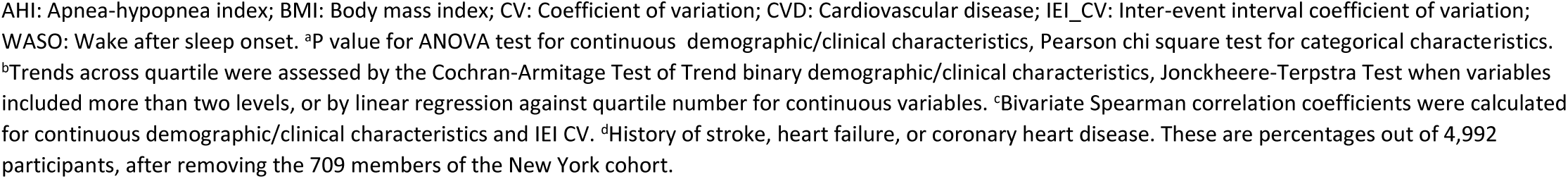
Demographic, anthropometric, and clinical characteristics of participants in the mortality dataset, by IEI_CV quartile. Mean (SD) or number (%).

|  | Q1<br>N=1,426<br>n (%) | Q2<br>N=1,425<br>n (%) | Q3<br>N=1,425<br>n (%) | Q4<br>N=1,425<br>n (%) | Quartile<br>Difference<br>P value<sup>a</sup> | Test of<br>Trend<sup>b</sup><br>P value | Correlation<br>Coefficient<sup>c</sup><br>(P value) |
| --- | --- | --- | --- | --- | --- | --- | --- |
| **IEI COEFFICIENT OF VARIATION** |  |  |  |  |  |  |  |
| Range | 0.42 - 1.86 | 1.86 - 2.24 | 2.24-2.65 | 2.65-5.85 |  |  |  |
| Mean (sd) | 1.55 (0.27) | 2.05 (0.11) | 2.43 (0.11) | 3.11 (0.43) |  |  |  |
| **DEMOGRAPHICS** |  |  |  |  |  |  |  |
| Age, years | 65.2 (11.6) | 63.6 (11.1) | 62.0 (10.7) | 62.3 (11.0) | <0.001 | <0.001 | -0.11 (<0.001) |
| Female | 694 (48.7%) | 797 (55.9%) | 776 (54.5%) | 719 (50.5%) | <0.001 | 0.51 |  |
| Race |  |  |  |  | 0.67 | 0.08 |  |
| White | 1,203 (84.4%) | 1,200 (84.2%) | 1,218 (85.5%) | 1,232 (86.5%) |  |  |  |
| Black | 129 (9.0%) | 132 (9.3%) | 118 (8.3%) | 108 (7.6%) |  |  |  |
| Other | 94 (6.6%) | 93 (6.5%) | 89 (6.2%) | 85 (6.0%) |  |  |  |
| Smoking Status |  |  |  |  | 0.16 | 0.39 |  |
| Current | 140 (9.8%) | 125 (8.8%) | 147 (10.3%) | 137 (9.6%) |  |  |  |
| Former | 648 (45.4%) | 613 (43.0%) | 581 (40.8%) | 633 (44.4%) |  |  |  |
| Never | 638 (44.7%) | 687 (48.2%) | 697 (48.9%) | 655 (46.0%) |  |  |  |
| **SLEEP CHARACTERISTICS** |  |  |  |  |  |  |  |
| AHI3, events/hour | 16.6 (20.8) | 11.0 (12.4) | 11.7 (11.5) | 15.8 (12.4) | <0.001 | 0.31 | 0.14 (<0.001) |
| AHI0, events/hour | 37.6 (27.0) | 29.9 (17.8) | 30.9 (15.5) | 36.2 (15.0) | <0.001 | 0.20 | 0.08 (<0.001) |
| Arousals/hour | 22.3 (13.832) | 18.1 (9.0) | 17.8 (9.1) | 18.7 (9.5) | <0.001 | <0.001 | -0.08 (<0.001) |
| Event duration, sec | 21.2 (5.5) | 21.2 (4.6) | 21.3 (4.7) | 21.4 (4.6) | 0.64 | 0.22 | 0.03 (0.030) |
| Sleep time, hours | 5.8 (1.2) | 6.0 (1.0) | 6.0 (1.0) | 6.0 (1.0) | <0.001 | <0.001 | 0.06 (<0.001) |
| WASO, minutes | 68.4 (49.3) | 61.5 (44.0) | 58.9 (40.8) | 57.7 (41.3) | <0.001 | <0.001 | -0.08 (<0.001) |
| % Sleep <90% sat | 5.0 (13.2) | 2.8 (9.0) | 2.6 (8.9) | 3.7 (9.5) | <0.001 | 0.002 | 0.07 (<0.001) |
| Delta Heart Rate, bpm | 7.0 (3.5) | 6.8 (2.8) | 7.1 (2.9) | 7.2 (3.0) | 0.003 | <0.001 | 0.07 (<0.001) |
| Hypoxic burden, desat% × h | 64.3 (86.4) | 42.3 (46.5) | 42.5 (38.6) | 52.9 (41.8) | <0.001 | <0.001 | 0.09 (<0.001) |
| OSA severity |  |  |  |  | <0.001 | <0.001 |  |
| None | 582 (40.8%) | 567 (39.8%) | 463 (32.5%) | 263 (18.5%) |  |  |  |
| Mild | 345 (24.2%) | 517 (36.3%) | 582 (40.8%) | 560 (39.3%) |  |  |  |
| Moderate | 217 (15.2%) | 226 (15.9%) | 285 (20.0%) | 425 (29.8%) |  |  |  |
| Severe | 282 (19.8%) | 115 (8.1%) | 95 (6.7%) | 177 (12.4%) |  |  |  |
| **CLINICAL CHARACTERISTICS** |  |  |  |  |  |  |  |
| BMI, kg/m<sup>2</sup> | 28.0 (5.2) | 27.8 (5.0) | 28.1 (4.8) | 28.8 (5.3) | <0.001 | <0.001 | 0.06 (<0.001) |
| Diabetes | 144 (10.1%) | 99 (6.9%) | 87 (6.1%) | 120 (8.4%) | <0.001 | 0.066 |  |
| Hypertension | 675 (47.3%) | 574 (40.3%) | 588 (41.3%) | 602 (42.2%) | <0.001 | 0.015 |  |
| Prevalent Heart Disease<sup>d</sup> | 200 (14%) | 153 (10.7%) | 131 (9.2%) | 135 (9.5%) | <0.001 | <0.001 |  |
| **OUTCOMES** |  |  |  |  |  |  |  |
| Deaths | 435 (30.5%) | 314 (22.0%) | 272 (19.1%) | 266 (18.7%) | <0.001 | <0.001 |  |
| Person-years of follow-up | 14,942 | 15,749 | 16,033 | 16,062 |  |  |  |
| Mortality rate per 1,000 person-years | 29.1 | 19.9 | 17.0 | 16.6 |  |  |  |

AHI: Apnea-hypopnea index; BMI: Body mass index; CV: Coefficient of variation; CVD: Cardiovascular disease; IEI\_CV: Inter-event interval coefficient of variation; WASO: Wake after sleep onset. <sup>a</sup>P value for ANOVA test for continuous demographic/clinical characteristics, Pearson chi square test for categorical characteristics. <sup>b</sup>Trends across quartile were assessed by the Cochran-Armitage Test of Trend binary demographic/clinical characteristics, Jonckheere-Terpstra Test when variables included more than two levels, or by linear regression against quartile number for continuous variables. <sup>c</sup>Bivariate Spearman correlation coefficients were calculated for continuous demographic/clinical characteristics and IEI CV. <sup>d</sup>History of stroke, heart failure, or coronary heart disease. These are percentages out of 4,992 participants, after removing the 709 members of the New York cohort.

### Analyses

Descriptive statistics were calculated for baseline characteristics and potential confounders of interest. Differences across quartiles of IEI_CV were evaluated using an analysis of variance (ANOVA) or Kruskal-Wallis test for continuous variables, and Pearson’s chi-squared test for categorical variables. Associations between IEI_CV and mortality, and between IEI_CV and incident CVD, were examined using Cox proportional hazards regression to estimate hazard ratios (HRs) and 95% confidence intervals (CIs). Survival time was defined as the time from baseline polysomnography to death or to the first CVD event, respectively. Participants who did not experience the outcome were censored at the time of their last known event-free status. Continuous variables were assessed for linearity with the log-hazard outcome and modeled as categorical variables if a non-linear association was observed. Models for both endpoints met the assumption of proportional hazards (Grambsch and Therneau test).^30^ Plots of scaled Schoenfeld residuals for each independent variable against four separate functions of time to event (identity, natural log, 1 - Kaplan-Meier product-limit estimate, and rank time scaling) showed no overt violation of the proportional hazards assumption.

Significance was set at α=0.05. Analyses were conducted in STATA 17.0 (StataCorp, College Station, TX).

## RESULTS

### Group characteristics

Participants in the two analytical subsets had similar demographic and anthropometric characteristics (**e-Table 2**). On average, participants were 63 years old and just over half female and about 85% self-identified as white. On average, OSA was categorized as mild but had a wide distribution (median AHI3 [IQR] = 8.9[3.6-18.5] and 8.5[3.4-17.7], for the mortality and prospective CVD subset, respectively. AHI0 was much higher (median[IQR] = 29.6[19.0-44.4] and 29.2[18.7-43.8], respectively). For all participants, the mean IEI (∼2 min) was approximately 60/AHI0 as expected.

**Table 2.** Cox proportional hazard ratios for all-cause mortality (top) or incident CVD (bottom). Adjusted for age, sex, BMI class, smoking status, race, OSA severity, and prevalent hypertension and diabetes. **\**P*<.05, ** *P*<.01, *** *P*<.001; #** *P*<.10.

|  |  | All-Cause Mortality |  |  |  |  |  |  |  |
| --- | --- | --- | --- | --- | --- | --- | --- | --- | --- |
|  |  | All |  | AHI0≥10 |  | AHI3≥5 |  | All + Max Sleep Bout Duration |  |
|  |  | HR | 95% CI | HR | 95% CI | HR | 95% CI | HR | 95% CI |
| IEI CV | Q1 (most regular) | **1.40\*\*\*** | **1.20,1.64** | **1.41\*\*\*** | **1.20,1.65** | **1.37\*\*\*** | **1.15,1.63** | **1.41\*\*\*** | **1.15,1.75** |
|  | Q2 | 1.13 | 0.96,1.33 | 1.13 | 0.95,1.33 | 1.06 | 0.88,1.28 | 1.14 | 0.94,1.38 |
|  | Q3 | 1.13 | 0.95,1.34 | 1.11 | 0.94,1.32 | 1.08 | 0.89,1.30 | 1.13 | 0.95,1.36 |
|  | Q4 (most variable) | 1 | 1.00,1.00 | 1 | 1.00,1.00 | 1 | 1.00,1.00 | 1 | 1.00,1.00 |
| # people |  | 5,701 |  | 5,333 |  | 3,826 |  | 5,701 |  |
|  |  | Incident CVD |  |  |  |  |  |  |  |
|  |  | All |  | AHI0>10 |  | AHI3≥5 |  | All + Max Sleep Bout Duration |  |
|  |  | HR | 95% CI | HR | 95% CI | HR | 95% CI | HR | 95% CI |
| IEI CV | Q1 (most regular) | **1.29\*** | **1.06,1.56** | **1.34\*\*** | **1.11,1.64** | **1.30\*** | **1.04,1.61** | 1.15 | 0.89,1.50 |
|  | Q2 | **1.22\*** | **1.00,1.49** | 1.22# | 1.00,1.49 | **1.25\*** | **1.00,1.55** | 1.13 | 0.90,1.43 |
|  | Q3 | 1.17 | 0.96,1.43 | 1.15 | 0.94,1.41 | 1.11 | 0.88,1.39 | 1.11 | 0.90,1.38 |
|  | Q4 (most variable) | 1 | 1.00,1.00 | 1 | 1.00,1.00 | 1 | 1.00,1.00 | 1 | 1.00,1.00 |
| # people |  | 4,373 |  | 4,083 |  | 2885 |  | 4,373 |  |

### IEI_CV: relationships with baseline characteristics

Within-participants, IEI_CV was approximately normally distributed, whereas the underlying distribution of individual inter-event intervals was approximately log-normal. Notably, individuals with comparable OSA severity, as defined by similar AHI3 values, exhibited markedly different nocturnal event patterns, resulting in substantial variation in IEI_CV (**Figure 2**). The IEI_CV distributions were similar in women and men (**e-Figure 1**).

**Figure 2.**
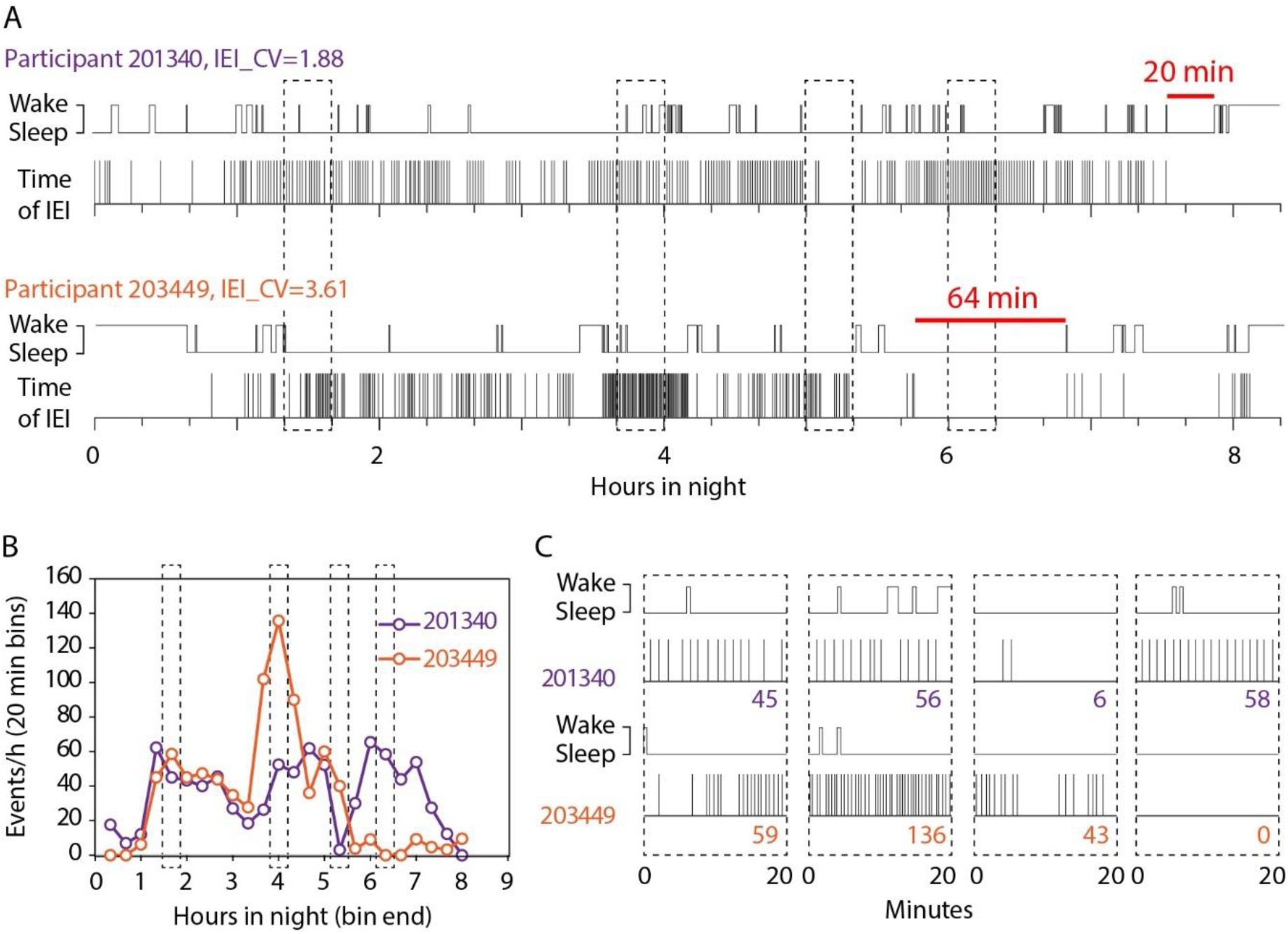
Event occurrence and changes in event frequency through the night. A) Eight hour sleep records for two SHHS subjects with AHI0=35.0. A simplified histogram shows wakefulness and sleep. The time of each respiratory event is shown as a vertical line. Participant 203449 (lower) has much greater inter-event variability (IEI_CV=3.61) and longer stretches of undisturbed sleep (maximum = 64 min, red line) compared to participant 201340 (IEI_CV=1.88, longest sleep = 20 min). B) Plot of serial AHI0 in 20 min bins showing higher peak AHI0 and long stretches of low AHI0 for participant 203449. C) Detail of 20 min boxes in A and B. Numbers in the panel are the AHI0 for that bin.

In the mortality analysis (n=5,701), bivariate analyses showed that IEI_CV was weakly but significantly correlated with several anthropometric and sleep characteristics (|ρ|’s from 0.03 to 0.14), **Table 1**); but overall, the differences across quartiles were small. Individuals in Q1 (i.e., most regularly occurring events) were older and had a higher proportion of individuals with diabetes and hypertension than individuals in Q2-4. Those in Q1 also had the highest proportion of individuals with both no OSA and severe OSA, and also exhibited polysomnographic findings of shorter total sleep time, longer wake after sleep onset (WASO), and a higher arousal index. Similar patterns of associations were observed in the CVD analytical subset (n=4,373; **e-Table 3**).

**Table 3.** Shorter maximum uninterrupted sleep durations are associated with an increased risk for mortality and incident CVD (adjusted as in Table 2). IEI_CV is not included in the model.

|  |  | Mortality |  |  | Incident CVD |  |  |
| --- | --- | --- | --- | --- | --- | --- | --- |
|  |  | Max sleep duration (range, min) | HR | ci95 | Max sleep duration (range, min) | HR | ci95 |
| Maximum IEI | Q1 <Shortest> | 0.70 – 21.0 | **1.20\*** | **1.01,1.43** | 0.90 – 21.6 | **1.40\*\*** | **1.12,1.76** |
|  | Q2 | 21.0 – 31.6 | 0.98 | 0.83,1.17 | 21.6 – 32.0 | **1.35\*\*** | **1.09,1.67** |
|  | Q3 | 31.6 – 43.0 | 0.90 | 0.76,1.08 | 32.0 – 43.4 | 1.13 | 0.90,1.41 |
|  | Q4 <Longest> | 43.0 – 140.0 | 1 | 1.00,1.00 | 43.4 – 113.6 | 1 | 1.00,1.00 |

In both the mortality and CVD analyses, IEI-CV had a U-shaped association with several OSA-related measures, including AHI3, AHI0, desaturation time below 90%, and hypoxic burden (i.e., higher in both Q1 and Q4 than in the middle two quartiles). This suggests that those with the most regular events have the lowest arousal thresholds and may stay awake longer once awoken. In contrast, ΔHR increased across quartiles of Q1 to Q4 (trend test, *P*<.001). Respiratory event duration did not differ across quartiles.

### Associations with all-cause mortality

In unadjusted analyses, people with more regular IEI, indicated by low IEI_CV (Q1), experienced greater mortality (Kaplan-Meier, *P*<.001, **Figure 3A**, **Table 1**). Overall, those who died had a left-shift in their IEI_CV distribution suggesting more regular event spacing (**Figure 3B**). In a Cox proportional hazard model adjusted for age, sex, BMI class, smoking status, race, AHI3, prevalent hypertension, and diabetes, participants in the lowest quartile of IEI_CV (Q1) had a 40% higher risk of all-cause mortality compared with those in the highest quartile (Q4) (HR[95%CI] = 1.40[1.20–1.64], *P*<.001; **Table 2**). To address the potential skewing of IEI_CV estimates in participants with few respiratory events, the analysis was repeated among individuals with at least 10 events per hour; results were materially unchanged (HR = 1.41[1.20–1.65], *P*<.001). Finally, restricting the analysis to participants with clinically defined mild or greater OSA (AHI3 ≥ 5) yielded similar findings, with Q1 still associated with an increased risk of all-cause mortality (HR = 1.37[1.15–1.63], *P*<.001; **Table 2**).

**Figure 3.**
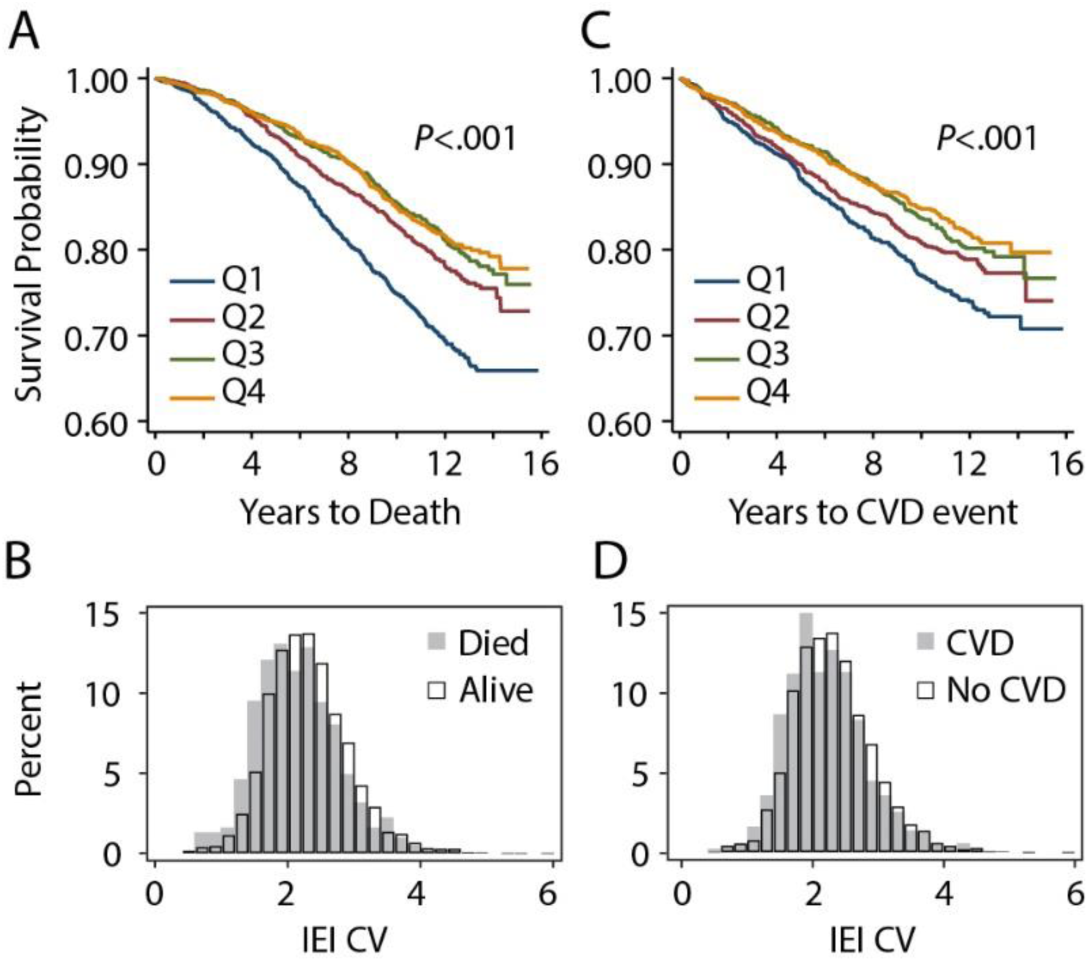
Kaplan-Meier survival curves by IEI_CV quartile, indicating earlier mortality (A) and incident cardiovascular disease events (C) in quartile 1 (lowest IEI_CV indicating least variability in inter-event interval). Kaplan-Meier p value indicates that at least two quartiles differ significantly. The overall distribution of mean inter-event interval is left-shifted in those who died (B, t-test, t_5699_ = 8.19, *P*<.001) or experienced a cardiovascular event (D, t_4990_=5.45, *P*<.001) compared to those that did not.

A sensitivity analysis was conducted for potential confounding or mediation by the presence of prevalent heart disease (not including the 709 participants of the New York cohort) and by other OSA metrics that have been associated with mortality, including obstructive event duration, hypoxic burden, and ΔHR.^8,9,29^ Including each singly, the hazard rate was largely unchanged from 1.40, ranging from 1.35 to 1.38. With all four combined (n=4,882), the hazard rate was only minimally reduced (Q1 HR=1.32[1.12-1.57], *P*=.001).

### Associations with incident CVD

People with low IEI_CV (Q1) experienced greater rates of incident CVD in unadjusted analyses (**Table 2**, **Figure 3C**). As above, the distribution of IEI_CV was shifted leftward for participants who experienced an incident CVD event (**Figure 3D**). Participants in both the lowest (Q1) and second (Q2) quartiles of IEI_CV exhibited a higher risk of incident CVD compared with those in the highest quartile, corresponding to the most variable inter-event intervals (HR = 1.29[1.06-1.56], *P*=.011 and HR = 1.22[1.00-1.49], *P*=.049, respectively). These associations were materially unchanged in sensitivity analyses restricted to participants with at least 10 respiratory events per hour and to those with clinically defined mild or greater OSA (**Table 2**).

As above, sensitivity analyses separately including event duration, hypoxic burden, and ΔHR categories did not alter the hazard rate (1.27-1.30). Considered together, the hazard rates were almost unchanged from 1.29 (Q1 HR=1.27[1.04, 1.55]).

### Stratification by sex

Both women and men had increased risk of mortality, with Q1 higher by 32% (*P*=.021) and 45% (*P*=.001), respectively (**e-Table 4**). For incident CVD, associations appear stronger for women (Q1 HR = 1.38[1.03-1.85]) than men (QI HR = 1.19[0.92-1.55]). However, a formal test for interaction of IEI_CV (continuous) by sex was non-significant (*P*=.144).

### Analyses of sleep bout duration

For each person, their longest bout of uninterrupted sleep ranged from 42 sec to 140 minutes. They were longest in people with both greater IEI variability and less severe OSA, while the shortest bouts occurred in people with both low IEI variability and severe OSA (**Figure 4A**). For both mortality and incident CVD, there was a right shift in the distribution towards longer maximum sleep intervals in those who did not experience a mortality or incident CVD outcome (**Figure 4B,C**). For mortality, adjustment of the primary Cox proportional hazards model for each person’s longest stretch of sleep did not appreciably influence the study findings (**Table 2**). For incident CVD, the effect estimate for the Q1 hazard ratio was reduced by 10.5% (from 1.29 to 1.15) and was no longer significant (*P*=.29) (**Table 2**). In exploratory hazard models based on each person’s longest sleep bout and independent of IEI_CV, we found that the those with the shortest (Q1) maximum sleep bout durations had significantly greater risk for both mortality (+20%, *P*=.043) and incident CVD (+40%, *P*=.003) (**Table 3**).

**Figure 4.**
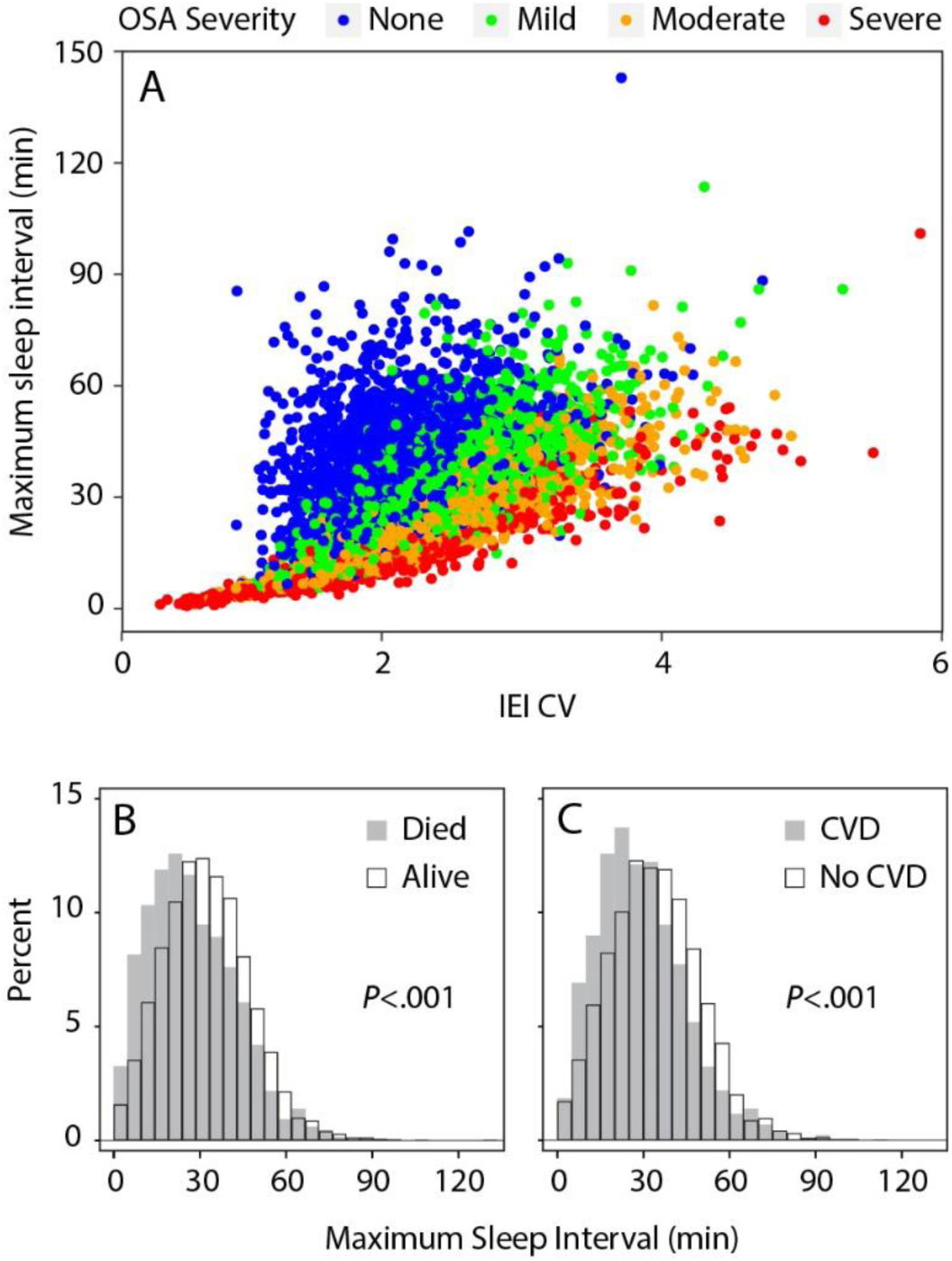
Relationships between IEI variability and maximum uninterrupted sleep bout. A) The longest undisturbed sleep interval increases with greater IEI variability and with less severe OSA. There is a pronounced tail on the left side of participants whose IEI CV is very low because the OSA is so severe. B and C) There is left shift towards shorter maximum sleep interval in those who died (B, t-test, t_5699_ = 10.63, *P*<.001) or experienced an incident CVD (C, t_4990_=10.62, *P*<.001).

## DISCUSSION

In this large, prospective community-based cohort, we identified a novel association between reduced inter-event variability—reflecting more regularly occurring obstructive respiratory events—and both all-cause mortality and incident cardiovascular disease. These associations remained robust after adjustment for multiple confounders and for established and emerging metrics of OSA severity, including the AHI, obstructive event duration, event-related heart rate response, and hypoxic burden. Together, these findings indicate that the physiological consequences of OSA are not determined solely by the frequency or cumulative burden of obstructive events, but are also influenced by their temporal organization across the night.

Individuals with lower inter-event variability were, on average, older and had a modestly higher prevalence of diabetes, hypertension, and cardiovascular disease, yet they spanned a wide range of OSA severity. Notably, measures of OSA severity demonstrated a U-shaped distribution across inter-event variability quartiles, with higher AHI, greater time spent with oxygen saturation below 90%, and increased hypoxic burden observed in both the lowest and highest quartiles. In contrast, indices of sleep fragmentation—including the arousal index, wake after sleep onset, and a measure of the longest uninterrupted sleep bout—were selectively worse among participants with the lowest inter-event variability. Taken together, these findings suggest that reduced inter-event variability characterizes a distinct sleep-disordered breathing phenotype that is predictive of adverse cardiovascular and mortality outcomes and captures clinically relevant information beyond that provided by the AHI.

Respiratory events can exhibit markedly different temporal patterns across the night—occurring in concentrated bursts or more regularly spaced intervals—thereby producing distinct profiles of uninterrupted sleep and clustering of physiological stressors.^31^ Those with the greatest IEI_CV will have periods of higher and lower event density. Periods of higher density may be deleterious by condensing sleep fragmentation, mild ischemia–reperfusion stress, intrathoracic pressure swings, and arousal-related autonomic activity and blood pressure surges.^32^ Periods of lower density may allow longer stretches of uninterrupted restorative sleep. Our results support the importance of the latter: the longest uninterrupted sleep bout in each person correlated with IEI_CV (**Figure 4**), independently predicted both mortality and incident CVD (**Table 3**), and attenuated the association between inter-event variability and incident CVD albeit without affecting the association with mortality (**Table 2**). The sleep bout duration may help explain evidence linking sleep fragmentation to incident CVD.^33–35^ The lack of an effect of including maximum bout duration on mortality likely reflects additional, more complex physiological processes.

It is plausible that low inter-event variability also reflects underlying disease processes characterized by reduced physiological signal complexity and impaired regulatory control.^36^ Analogous phenomena have been described in other systems, including the stereotyped breathing patterns of Cheyne–Stokes respiration, increased regularity of sleep arousals in hypertension,^24^ and higher intensive care unit mortality associated with reduced complexity in temporal body temperature.^37^ Decreased heart rate variability is another marker of cardiovascular risk, although interpretation of standard metrics may be challenging. Recently, measures of sino-atrial variability referred to as “heart rate fragmentation” were developed from overnight data and shown to be positively associated with older age, cognitive impairment, and atrial fibrillation.^38,39^ Changes in heart rate fragmentation from wakefulness to sleep have been proposed as a novel index of cardiac neuroautonomic renewability, with smaller decreases associated with poorer sleep quality and higher risk of CVD.^40^ Together, these studies highlight the importance of studying the dynamics of physiological signals during sleep, with various measures of respiratory and cardiac variability providing information on neuroautonomic control.

Our approach to measuring inter-event variability was built on the hypothesis that temporal sequencing of events during sleep could better reflect ongoing physiological processes. There have been recent efforts to predict obstructive events from physiological signals and how best to understand the source of variability in the respiratory event train. Chen et al.^31^ developed a model to predict instantaneous probability of a respiratory event occurring to improve patient phenotyping. This method can accommodate ordered to random to grouped event occurrences, even in people with the same number or events. Others have developed methods to predict apneas and hypopneas from various physiological data during sleep.^41,42^ A better understanding of how a given event and the participant’s physiology influence the probability of future events could point to specific mechanisms that relate to long term morbidity.

There has been growing interest in improved OSA phenotyping – including developing metrics that capture risk in both men and women.^6^ Mortality risk was observed in both sexes; in contrast, IEI_CV was a predictor of incident CVD only in women. This follows the pattern that we previously reported for event duration,^8^ and contrasts with some studies that reported a relationship of the AHI with mortality and cardiovascular risk only for men.^27,28,43,44^

Study strengths included the large and geographically diverse sample, rigorous assessment of cardiovascular disease endpoints and mortality outcomes, and standardized polysomnography and validated sleep scoring. A limitation of this study-as common for almost all epidemiological studies of OSA-is the reliance on a single night assessment. Misclassification however would likely bias results to the null. However, further research should assess the night-to-night reproducibility of the temporal structure of OSA. Finally, SHHS is the largest prospective research cohort of men and women with both standardized polysomnography and event adjudication, allowing us to test the predictive value of this metric. There is a need to replicate the findings in independent samples with similar data, including clinic-based samples where the severity of OSA and related symptoms is greater than in community samples.

This study demonstrates that the temporal distribution of obstructive respiratory events across the night holds prognostic information for future cardiovascular disease and mortality, with particularly strong associations for cardiovascular disease prediction in women. The combined evidence that high inter-event variability is associated with greater stretches of undisturbed sleep, that maximum undisturbed sleep interval was related to both endpoints, and that maximum sleep interval explained a portion of the risk for cardiovascular disease together point to the potential restorative role of sustained sleep periods and the deleterious impact of continuous sleep fragmentation. These results highlight the clinical relevance of nocturnal event timing and sleep continuity as physiologically meaningful dimensions of OSA that extend beyond many existing summary measures of disease severity.

## Data Availability

All data produced in the present study are available upon reasonable request to the authors.

## Acknowledgements

The authors thank Jeffery Emch for coding assistance, the Sleep Heart Health Study Reading Center team for expert sleep scoring, and the National Sleep Research Resource for providing the data.

## Conflicts of Interest

SR has received consulting fees from Amgen Inc and a stipend from the National Sleep Foundation in her role as editor of Sleep Health. SR’s institution received a contract from Google, Inc. AA serves as a consultant for Zoll Respicardia, Inspire, Cerebra, Eli Lilly, Amgen, and Apnimed. AA serves as an advisory board member for Incannex. Apnimed is developing pharmacological treatments for Obstructive Sleep Apnea. AA also reports a pending patent related to endo-phenotyping sleep apnea. AA’s interests were reviewed by Brigham and Women’s Hospital and Mass General Brigham in accordance with their institutional policies.

## Funding Information

NIH R21HL140377 (MPB, SR), R01HD109477 (MPB), R35HL135818, R24HL114473 (SR); American Sleep Medicine Foundation (ASMF) Focused Project Award (MPB), and the Oregon Institute of Occupational Health Sciences via funds from the Division of Consumer and Business Services of the State of Oregon (ORS 656.630).

## Contribution acknowledgement

AJS, SR, and MPB designed the research, analyzed data, and drafted and reviewed the manuscript. AA provided data and reviewed the analysis and the manuscript. FK and KAL provided analytical tools and reviewed the manuscript. SR oversaw data collection and made it available via the National Sleep Research Resource. All authors approved the final version and agree to be accountable for all aspects of the work. MPB is the guarantor of the content of the manuscript.

## Abbreviation List

AHI: apnea hypopnea index
CI: 95% confidence interval
HR: hazard ratio
IQR: interquartile range
NSRR: National Sleep Research Resource
OSA: obstructive sleep apnea
SHHS: Sleep Heart Health Study
IEI: Inter-event interval
IEI_CV: Inter-event interval coefficient of variation
CVD: cardiovascular disease
Q#: quartile #

## Notes

### Author Declarations

The Institutional Review Board of Oregon Health & Science University reviewed the study and waived ethical approval for this work as it is analysis of deidentified data.

## REFERENCES

1. Javaheri S, Barbe F, Campos-Rodriguez F, et al. Sleep apnea: types, mechanisms, and clinical cardiovascular consequences. J. Am. Coll. Cardiol. 2017;69(7):841–858.

2. Yeghiazarians Y, Jneid H, Tietjens JR, et al. Obstructive Sleep Apnea and Cardiovascular Disease: A Scientific Statement From the American Heart Association. Circulation. 2021;144(3):e56–e67.

3. Shahar E. Apnea-hypopnea index: time to wake up. Nat Sci Sleep. 2014;6:51–56.

4. Malhotra A, Ayappa I, Ayas N, et al. Metrics of sleep apnea severity: beyond the apnea-hypopnea index. Sleep. 2021;44(7).

5. Malhotra A, Mesarwi O, Pepin JL, Owens RL. Endotypes and phenotypes in obstructive sleep apnea. Curr. Opin. Pulm. Med. 2020;26(6):609–614.

6. Redline S, Azarbarzin A, Peker Y. Obstructive sleep apnoea heterogeneity and cardiovascular disease. Nat. Rev. Cardiol. 2023;20(8):560–573.

7. Borker PV, Reid M, Sofer T, et al. NREM apnea and hypopnea duration varies across population groups and physiologic traits. Am. J. Respir. Crit. Care Med. 2020:doi: 10.1164/rccm.202005-201808OC. Online ahead of print.

8. Butler MP, Emch JT, Rueschman M, et al. Apnea-hypopnea event duration predicts mortality in men and women in the Sleep Heart Health Study. Am. J. Respir. Crit. Care Med. 2019;199(7):903–912.

9. Azarbarzin A, Sands SA, Stone KL, et al. The hypoxic burden of sleep apnoea predicts cardiovascular disease-related mortality: the Osteoporotic Fractures in Men Study and the Sleep Heart Health Study. Eur. Heart J. 2018.

10. Kulkas A, Tiihonen P, Eskola K, Julkunen P, Mervaala E, Toyras J. Novel parameters for evaluating severity of sleep disordered breathing and for supporting diagnosis of sleep apnea-hypopnea syndrome. J. Med. Eng. Technol. 2013;37(2):135–143.

11. Muraja-Murro A, Kulkas A, Hiltunen M, et al. The severity of individual obstruction events is related to increased mortality rate in severe obstructive sleep apnea. J. Sleep Res. 2013.

12. Douglas NJ, White DP, Pickett CK, Weil JV, Zwillich CW. Respiration during sleep in normal man. Thorax. 1982;37(11):840–844.

13. Ferris BG, Jr., Pollard DS. Effect of deep and quiet breathing on pulmonary compliance in man. J. Clin. Invest. 1960;39(1):143–149.

14. Bartlett D, Jr. Origin and regulation of spontaneous deep breaths. Respir. Physiol. 1971;12(2):230–238.

15. Bendixen HH, Smith GM, Mead J. Pattern of Ventilation in Young Adults. J. Appl. Physiol. 1964;19:195–198.

16. Kubin L. Breathing during sleep. Handb. Clin. Neurol. 2022;188:179–199.

17. Butler MP, Smales C, Wu H, et al. The circadian system contributes to apnea lengthening across the night in obstructive sleep apnea. Sleep. 2015;38(11):1793–1801.

18. Ravesloot MJL, White D, Heinzer R, Oksenberg A, Pepin JL. Efficacy of the New Generation of Devices for Positional Therapy for Patients With Positional Obstructive Sleep Apnea: A Systematic Review of the Literature and Meta-Analysis. J. Clin. Sleep Med. 2017;13(6):813–824.

19. Charbonneau M, Marin JM, Olha A, Kimoff RJ, Levy RD, Cosio MG. Changes in obstructive sleep apnea characteristics through the night. Chest. 1994;106(6):1695–1701.

20. McSharry DG, Saboisky JP, Deyoung P, et al. A mechanism for upper airway stability during slow wave sleep. Sleep. 2013;36(4):555–563.

21. Haba-Rubio J, Janssens JP, Rochat T, Sforza E. Rapid eye movement-related disordered breathing: clinical and polysomnographic features. Chest. 2005;128(5):3350–3357.

22. Oksenberg A, Leppanen T. Duration of respiratory events in obstructive sleep apnea: Factors influencing the duration of respiratory events. Sleep Med. Rev. 2023;68:101729.

23. Jarczok MN, Weimer K, Braun C, et al. Heart rate variability in the prediction of mortality: A systematic review and meta-analysis of healthy and patient populations. Neurosci. Biobehav. Rev. 2022;143:104907.

24. Jamasebi R, Redline S, Patel SR, Loparo KA. Entropy-based measures of EEG arousals as biomarkers for sleep dynamics: applications to hypertension. Sleep. 2008;31(7):935–943.

25. Quan SF, Howard BV, Iber C, et al. The Sleep Heart Health Study: design, rationale, and methods. Sleep. 1997;20(12):1077–1085.

26. Zhang GQ, Cui L, Mueller R, et al. The National Sleep Research Resource: towards a sleep data commons. J. Am. Med. Inform. Assoc. 2018;25(10):1351–1358.

27. Punjabi NM, Caffo BS, Goodwin JL, et al. Sleep-disordered breathing and mortality: a prospective cohort study. PLoS Med. 2009;6(8):e1000132.

28. Gottlieb DJ, Yenokyan G, Newman AB, et al. Prospective study of obstructive sleep apnea and incident coronary heart disease and heart failure: the sleep heart health study. Circulation. 2010;122(4):352–360.

29. Azarbarzin A, Sands SA, Younes M, et al. The Sleep Apnea-Specific Pulse-Rate Response Predicts Cardiovascular Morbidity and Mortality. Am. J. Respir. Crit. Care Med. 2021;203(12):1546–1555.

30. Grambsch P, Therneau T. Proportional hazards tests and diagnostics based on weighted residuals Biometrika. 1994;81:515–526.

31. Chen S, Redline S, Eden UT, Prerau MJ. Dynamic models of obstructive sleep apnea provide robust prediction of respiratory event timing and a statistical framework for phenotype exploration. Sleep. 2022;45(12).

32. Somers VK, White DP, Amin R, et al. Sleep apnea and cardiovascular disease: an American Heart Association/American College Of Cardiology Foundation Scientific Statement from the American Heart Association Council for High Blood Pressure Research Professional Education Committee, Council on Clinical Cardiology, Stroke Council, and Council On Cardiovascular Nursing. In collaboration with the National Heart, Lung, and Blood Institute National Center on Sleep Disorders Research (National Institutes of Health). Circulation. 2008;118(10):1080–1111.

33. Badran M, Puech C, Gozal D. The cardiovascular consequences of chronic sleep fragmentation: Evidence from experimental models of obstructive sleep apnea. Sleep Med. 2025;132:106566.

34. Martynowicz H, Wichniak A, Wieckiewicz M. Sleep disorders and cardiovascular risk: Focusing on sleep fragmentation. Dent Med Probl. 2024;61(4):475–477.

35. Lutsey PL, McClelland RL, Duprez D, et al. Objectively measured sleep characteristics and prevalence of coronary artery calcification: the Multi-Ethnic Study of Atherosclerosis Sleep study. Thorax. 2015;70(9):880–887.

36. Goldberger AL, Amaral LA, Hausdorff JM, Ivanov P, Peng CK, Stanley HE. Fractal dynamics in physiology: alterations with disease and aging. Proc. Natl. Acad. Sci. U. S. A. 2002;99 Suppl 1:2466–2472.

37. Varela M, Churruca J, Gonzalez A, Martin A, Ode J, Galdos P. Temperature curve complexity predicts survival in critically ill patients. Am. J. Respir. Crit. Care Med. 2006;174(3):290–298.

38. Costa MD, Davis RB, Goldberger AL. Heart Rate Fragmentation: A New Approach to the Analysis of Cardiac Interbeat Interval Dynamics. Front. Physiol. 2017;8:255.

39. Costa MD, Redline S, Hughes TM, Heckbert SR, Goldberger AL. Prediction of Cognitive Decline Using Heart Rate Fragmentation Analysis: The Multi-Ethnic Study of Atherosclerosis. Front. Aging Neurosci. 2021;13:708130.

40. Costa MD, Heckbert SR, Redline S, Goldberger AL. Method to quantify the impact of sleep on cardiac neuroautonomic functionality: application to the prediction of cardiovascular events in the Multi-Ethnic Study of Atherosclerosis. Am. J. Physiol. Regul. Integr. Comp. Physiol. 2022;323(6):R968–R978.

41. Chen Y, Yang S, Li H, Wang L, Wang B. Prediction of Sleep Apnea Events Using a CNN-Transformer Network and Contactless Breathing Vibration Signals. Bioengineering (Basel). 2023;10(7).

42. Waxman JA, Graupe D, Carley DW. Automated prediction of apnea and hypopnea, using a LAMSTAR artificial neural network. Am. J. Respir. Crit. Care Med. 2010;181(7):727–733.

43. Redline S, Yenokyan G, Gottlieb DJ, et al. Obstructive sleep apnea-hypopnea and incident stroke: the Sleep Heart Health Study. Am. J. Respir. Crit. Care Med. 2010;182(2):269–277.

44. Yeboah J, Redline S, Johnson C, et al. Association between sleep apnea, snoring, incident cardiovascular events and all-cause mortality in an adult population: MESA. Atherosclerosis. 2011;219(2):963–968.

