## Supplementary material for "Regularity in occurrence of respiratory-related events in sleep predicts cardiovascular disease and mortality": e-Table 4

**e-Table 4.** Hazard ratios for mortality (top) and incident CVD (bottom) by IEI_CV quartile and stratified by sex. Quartiles were not re-leveled. Adjustments as in Table 2. ****P*<.05, ***P*<.01, ****P*<.001**, #*P*<.10

|  |  | Mortality | | | | | |
| --- | --- | --- | --- | --- | --- | --- | --- |
|  |  | Women | | | Men | | |
|  |  | N | HR | ci95 | N | HR | ci95 |
| IEI_CV | Q1 | **694** | **1.32*** | **1.04,1.68** | **732** | **1.45***** | **1.17,1.79** |
|  | Q2 | 797 | 1.23# | 0.97,1.56 | 628 | 1.04 | 0.82,1.32 |
|  | Q3 | 776 | 1.04 | 0.81,1.34 | 649 | 1.21 | 0.96,1.54 |
|  | Q4 | 719 | 1 | 1.00,1.00 | 706 | 1 | 1.00,1.00 |
|  |  | CVD | | | | | |
|  |  | N | HR | ci95 | N | HR | ci95 |
| IEI_CV | Q1 | 572 | **1.38*** | **1.03,1.85** | 522 | 1.19 | 0.92,1.55 |
|  | Q2 | 644 | 1.25 | 0.93,1.67 | 449 | 1.20 | 0.91,1.58 |
|  | Q3 | 624 | 1.05 | 0.77,1.41 | 469 | 1.26# | 0.96,1.66 |
|  | Q4 | 600 | 1 | 1.00,1.00 | 493 | 1 | 1.00,1.00 |
