## Supplementary material for "Regularity in occurrence of respiratory-related events in sleep predicts cardiovascular disease and mortality": e-Table 3

**e-Table 3.** Demographic, anthropometric, and clinical characteristics of participants in the incident CVD dataset, by IEI_CV quartile**.** Mean (SD) or number (%).

|  | **Q1** | **Q2** | **Q3** | **Q4** | **Quartile Difference** *P* value^a^ | **Test of Trend**^b^ *P* value | **Correlation Coefficient**^c^ (*P* value) |
| --- | --- | --- | --- | --- | --- | --- | --- |
|  | N=1,094 | N=1,093 | N=1,093 | N=1,093 |  |  |  |
|  | n (%) | n (%) | n (%) | n (%) |  |  |  |
| **IEI COEFFICIENT OF VARIATION** |  |  |  |  |  |  |  |
| Range | 0.42 - 1.86 | 1.86 - 2.24 | 2.24-2.65 | 2.65-5.85 |  |  |  |
| Mean (sd) | 1.57 (0.26) | 2.05 (0.11) | 2.43 (0.11) | 3.11 (0.44) |  |  |  |
| **DEMOGRAPHICS** |  |  |  |  |  |  |  |
| Age, years | 64.8 (11.5) | 63.5 (10.8) | 62.0 (10.6) | 62.2 (10.8) | <0.001 | <0.001 | -0.10 (<0.001) |
| Female | 572 (52.3%) | 644 (58.9%) | 624 (57.1%) | 600 (54.9%) | 0.012 | 0.37 |  |
| Race |  |  |  |  | 0.56 | 0.08 |  |
| White | 939 (85.8%) | 942 (86.2%) | 957 (87.6%) | 961 (87.9%) |  |  |  |
| Black | 71 (6.5%) | 76 (7.0%) | 67 (6.1%) | 71 (6.5%) |  |  |  |
| Other | 84 (7.7%) | 75 (6.9%) | 69 (6.3%) | 61 (5.6%) |  |  |  |
| Smoking Status |  |  |  |  | 0.25 | 0.81 |  |
| Current | 110 (10.1%) | 99 (9.1%) | 119 (10.9%) | 102 (9.3%) |  |  |  |
| Former | 481 (44.0%) | 471 (43.1%) | 434 (39.7%) | 488 (44.6%) |  |  |  |
| Never | 503 (46.0%) | 523 (47.8%) | 540 (49.4%) | 503 (46.0%) |  |  |  |
| **SLEEP CHARACTERISTICS** |  |  |  |  |  |  |  |
| AHI3, events/hour | 15.4 (19.9) | 10.7 (12.4) | 11.3 (11.0) | 15.6 (12.1) | <0.001 | 0.60 | 0.17 (<0.001) |
| AHI0, events/hour | 36.4 (26.8) | 29.5 (18.1) | 30.7 (15.3) | 36.3 (14.9) | <0.001 | 0.77 | 0.11 (<0.001) |
| Arousals/hour | 21.7 (12.9) | 18.0 (8.7) | 17.8 (9.1) | 18.7 (9.5) | <0.001 | <0.001 | -0.07 (<0.001) |
| Event duration, sec | 20.9 (5.1) | 21.1 (4.5) | 21.1 (4.5) | 21.2 (4.5) | 0.33 | 0.087 | 0.04 (0.009) |
| Sleep time, hours | 5.9 (1.1) | 6.1 (1.0) | 6.0 (1.0) | 6.1 (1.0) | <0.001 | 0.006 | 0.05 (0.010) |
| WASO, minutes | 66.2 (47.2) | 60.7 (42.7) | 59.3 (41.2) | 57.8 (40.7) | <0.001 | <0.001 | -0.06 (0.001) |
| % Sleep <90% sat | 4.4 (12.0) | 2.6 (8.6) | 2.5 (8.8) | 3.6 (9.2) | <0.001 | 0.057 | 0.08 (<0.001) |
| Delta Heart Rate, bpm | 6.9 (3.1) | 6.7 (2.5) | 7.0 (2.6) | 7.2 (2.8) | <0.001 | 0.001 | 0.08 (<0.001) |
| Hypox burden, desat% × h | 58.1 (73.2) | 39.1 (39.7) | 40.0 (34.1) | 51.1 (38.5) | <0.001 | <0.001 | 0.11 (<0.001) |
| OSA severity |  |  |  |  | <0.001 |  |  |
| None | 472 (43.1%) | 449 (41.1%) | 363 (33.2%) | 204 (18.7%) |  |  |  |
| Mild | 268 (24.5%) | 395 (36.1%) | 454 (41.5%) | 435 (39.8%) |  |  |  |
| Moderate | 160 (14.6%) | 163 (14.9%) | 216 (19.8%) | 314 (28.7%) |  |  |  |
| Severe | 194 (17.7%) | 86 (7.9%) | 60 (5.5%) | 140 (12.8%) |  |  |  |
| **CLINICAL CHARACTERISTICS** |  |  |  |  |  |  |  |
| BMI, kg/m^2^ | 28.1 (5.2) | 28.0 (5.0) | 28.1 (4.7) | 28.9 (5.3) | <0.001 | <0.001 | 0.06 (<0.001) |
| Diabetes | 97 (8.9%) | 69 (6.3%) | 55 (5.0%) | 87 (8.0%) | 0.002 | 0.25 |  |
| Hypertention | 439 (40.1%) | 355 (32.5%) | 369 (33.8%) | 390 (35.7%) | 0.001 | 0.062 |  |
| Prevalent Heart Disease | 0 (0%) | 0 (0%) | 0 (0%) | 0 (0%) |  |  |  |
| **OUTCOMES** |  |  |  |  |  |  |  |
| Incident CVD | 262 (23.9%) | 220 (20.1%) | 199 (18.2%) | 186 (17.0%) | <0.001 | <0.001 |  |
| Person-years of follow-up | 11,524 | 12,034 | 12,153 | 12,201 |  |  |  |
| CVD rate / 1,000 person-years | 22.7 | 18.3 | 16.4 | 15.2 |  |  |  |

AHI: Apnea-hypopnea index; BMI: Body mass index; CV: Coefficient of variation; CVD: Cardiovascular disease; IEI_CV: Inter-event interval coefficient of variation; WASO: Wake after sleep onset. ^a^P value for ANOVA test for continuous demographic/clinical characteristics, Pearson chi square test for categorical characteristics. ^b^Trends across quartile were assessed by the Cochran‐Armitage Test of Trend binary demographic/clinical characteristics, Jonckheere-Terpstra Test when variables included more than two levels, or by linear regression against quartile number for continuous variables. ^c^Bivariate Spearman correlation coefficients were calculated for continuous demographic/clinical characteristics and IEI CV.
