## Supplementary material for "Regularity in occurrence of respiratory-related events in sleep predicts cardiovascular disease and mortality": e-Table 2

**e-Table 2.** Selected baseline characteristics Mean (SD) [Range] or N or %.

|  | **All-cause mortality dataset** | **Incident CVD dataset** |
| --- | --- | --- |
| **N** | 5701 | 4373 |
| **Age, yr** | 63.30 (11.17) [39-90] | 63.13 (10.99) [39-90] |
| **BMI, kg/m^2^** | 28.16 (5.09) [18-50] | 28.29 (5.05) [18-50] |
| **Female sex, %** | 52.4 | 55.8 |
| **Race, %** |  |  |
| **White** | 85.1 | 86.9 |
| **Black** | 8.5 | 6.5 |
| **Other** | 6.3 | 6.6 |
| **Smoking Status, %** |  |  |
| **Current** | 9.6 | 9.8 |
| **Former** | 43.4 | 42.9 |
| **Never** | 47.0 | 47.3 |
| **AHI3, events/h** | 13.8 (15.0) [0-157] | 13.2 (14.5) [0-157] |
| **AHI0, events/h** | 33.7 (19.7) [0-173] | 33.2 (19.6) [1-173] |
| **IEI mean, sec** | 116 (91) [9-1516] | 118 (89) [8-1018] |
| **IEI count, intervals/night** | 213 (115) [14-944] | 213 (116) [15-944] |
| **OSA severity, %** |  |  |
| **None** | 32.9 | 34.0 |
| **Mild** | 35.2 | 35.5 |
| **Moderate** | 20.2 | 19.5 |
| **Severe** | 11.7 | 11.0 |
| **Prevalent Disease, %** |  |  |
| **Diabetes** | 7.9 | 7.0 |
| **Hypertension** | 42.8 | 35.5 |
| **Coronary Heart Disease** | 8.8† | 0 |
| **Stroke** | 3.1† | 0 |
| **Heart Failure** | 2.8† | 0 |
| **Prevalent CVD** | 12.4† | 0 |
| **OUTCOMES** |  |  |
| **Deaths** | 1287 | 859* |
| **Incident CVD** | 1190*† | 867 |
| **Person-years of follow-up** | 62,785 | 47,912 |

AHI: Apnea-hypopnea index; BMI: Body mass index; CV: Coefficient of variation; CVD: Any cardiovascular disease; IEI: Inter-event interval IEI_CV: IEI coefficient of variation; WASO: Wake after sleep onset. Data are presented as number, percent, or mean (sd) [range]. * Not used as endpoints. † The 709 participants of the New York cohort are not included in these numbers, so the percentage is of a denominator of 4,992 people.
