## Supplementary material for "Regularity in occurrence of respiratory-related events in sleep predicts cardiovascular disease and mortality": e-Table 1

**e-Table 1.** Variables used in analyses. The NSRR ID is the variable name used by NSRR. as coded by the National Sleep Research Resource database.

| **Data Category** | **Variable** | **NSRR ID** | **Other source** |
| --- | --- | --- | --- |
| Baseline demographic data and sleep and clinical covariates | Age | age_s1 |  |
|  | Sex | nsrr_sex |  |
|  | BMI | bmi_s1 |  |
|  | Race | race |  |
|  | Smoking status | smokstat_s1 |  |
|  | Sleep Characteristics |  |  |
|  | Total sleep duration | slp_rdi (now slpprdp) |  |
|  | Wake after sleep onset (WASO) | WASO |  |
|  | Percent sleep time below 90% sat | pctlt90 (now pctsa90h) |  |
|  | Event_Count |  | Number of apneas and hypopneas during sleep in XML record. |
|  | Arousal index | ai_all |  |
|  | AHI3 (AHI with 3% desaturation) | rdi3p |  |
|  | AHI0 (AHI, any desaturation) |  | 60*Event_count/slp_rdi |
|  | Obstructive event duration |  | Butler et al., 2019 |
|  | Event-specific heart rate increase |  | Azarbarzin et al., 2021 |
|  | Hypoxic burden |  | Azarbarzin et al., 2018 |
|  | Hypertension | HTNDerv_s1 |  |
|  | Diabetes, defined by any one of: |  |  |
|  | Self-report, | ParRptDiab |  |
|  | Use of insulin, or | INSULN1 |  |
|  | Use of oral hypoglycemic agent | OHGA1 |  |
|  | Parent Study | parentstudy | NSRR special request |
| Prevalent cardiovascular disease | Congestive heart failure | prev_chf |  |
|  | Stroke | prev_stk |  |
|  | Coronary heart disease, defined by: |  |  |
|  | Myocardial infarction (MI) | prev_mi |  |
|  | Procedures related to MI | prev_mip |  |
|  | Revascularization procedures | prev_revpro |  |
| All-cause mortality | Vital status at censor data | vital |  |
|  | Time to last contact or death | censdate |  |
| Incident cardiovascular disease | Any CVD event since baseline | any_cvd |  |
|  | Time to last contact or CVD event, defined as the minimum time from baseline to the following: |  |  |
|  | Last contact or death | censdate |  |
|  | First myocardial infarction | mi_date |  |
|  | Procedure related to MI | mip_date |  |
|  | Stroke | stk_date |  |
|  | Fatal coronary heart event | chd_dthdt |  |
|  | Fatal cardiovascular event | cvd_dthdt |  |
|  | Congestive heart failure | chf_date |  |
|  | Revascularization procedure | revpro_date |  |
|  | PTCA | ptca_date |  |
|  | CABG | cabg_date |  |

Azarbarzin A, Sands SA, Stone KL, et al. The hypoxic burden of sleep apnoea predicts cardiovascular disease-related mortality: the Osteoporotic Fractures in Men Study and the Sleep Heart Health Study. *Eur. Heart J.* 2018.

Azarbarzin A, Sands SA, Younes M, et al. The Sleep Apnea-Specific Pulse-Rate Response Predicts Cardiovascular Morbidity and Mortality. *Am. J. Respir. Crit. Care Med.* 2021;203(12):1546-1555.

Butler MP, Emch JT, Rueschman M, et al. Apnea-hypopnea event duration predicts mortality in men and women in the Sleep Heart Health Study. *Am. J. Respir. Crit. Care Med*. 2019;199(7):903-912.
