## Supplementary material for "Regularity in occurrence of respiratory-related events in sleep predicts cardiovascular disease and mortality": e-Figure 1

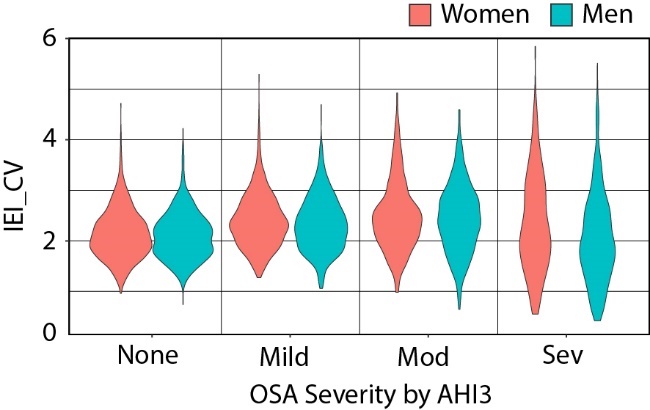

**e-Figure 1**. IEI_CV distributions are shown as violin plots for women and men at multiple OSA severity levels.
